# Efficacy and Safety of Guduchi Ghan Vati in the Management of Asymptomatic COVID-19 Infection: An Open Label Feasibility Study

**DOI:** 10.1101/2020.09.20.20198515

**Authors:** Abhimanyu Kumar, Govind Prasad, Sanjay Srivastav, Vinod Kumar Gautam, Neha Sharma

## Abstract

**Background:** Guduchi Ghan Vati (aqueous extract of *Tinospora cordifolia*) is an essential herbal plant in Indian traditional medicine (Ayurveda) that is well documented as an immunomodulator and antimicrobial agent. A recent in silico study found the therapeutic efficacy of Guduchi against SARS-CoV-2. Based on available evidence, we conducted a feasibility study of the safety and efficacy of Guduchi Ghan Vati in asymptomatic patients with covid-19.

**Patients and methods:** An open label, feasibility trial was conducted on 46 patients in the hospital setting. A single-arm study with no control group and blinding was executed in Jodhpur, Rajasthan, India. All patients orally received 2 tablets (1000 mg) twice daily for 2 weeks. Clinical parameters were collected at baseline, day 3, day 7 and day 14. Patients were continuously monitored for side effects and adverse reactions during the study period..

**Results:** Out of 46 asymptomatic patients included in the study, 40 completed the 14-day follow-up period. None developed any Covid-19 symptoms after admission to the hospital. On day 3 post-treatment, viral clearance was reported in 16 (32.5%) patients. By the end of D-7, 38 (95%) patients had viral load disappearance. Follow-up at D-14 showed that all participants tested negative.

**Conclusion:** In adult patients with asymptomatic Covid-19, Gudhuchi Ghan Vati could be effective. Randomized controlled trials with larger sample sizes in patients with Covid-19 are urgently needed to confirm the definite benefit with Ayurveda.

## Introduction

Severe acute respiratory syndrome coronavirus 2 (SARS-CoV-2) is a constantly evolving threat with over 25.8 million active cases and over 859 K deaths worldwide.^1^ Almost 80% of Covid-19 confirmed patients suffer from mild to moderate symptoms and recover on their own. Among these large proportion of low-risk Covid-19 cases, asymptomatic patients are merging higher. Asymptomatic patients appear with no clinical symptoms but are contagious.^2^

The emergence of asymptomatic patients poses a significant challenge to the prevention and treatment of the epidemic. Most clinical drugs and supportive therapies are focused on severe patients, whereas treatment options for asymptomatic to mild symptoms remain unclear. Asymptomatic to symptomatic cases may evolve to the most severe disease form without treatment indications. Standard guidelines suggest isolation for asymptomatic cases to control the spread of virus.^3^ There have not been any treatment options that reduce the viral load or preventive options that reduce the risk of developing severe conditions.

Various medicinal plants in Ayurveda (Indian Traditional Medicine)^4^ have been valued as immunomodulators,^5,6^ antiviral^7,8^ and antimicrobials.^9-12^ These properties have been scientifically investigated previously with promising results. Among them, Tinospora Cordifolia (Guduchi/ Giloy),^13^ is a unique Ayurvedic classical preparation called Guduchi Ghav Vati. With its pharmacological functions and medicinal values described, applications of Guduchi in countering various disorders are very common. Its usage as an immunomodulator,^14,15^ antioxidant,^16^ anti-microbial^17^ and anti-cancer properties,^18,19^ made it an interesting focus for Covid-19 management. Recently, an in silico pharmacology study^20^ showed the feasible efficacy of Guduchi against coronavirus in general and SARS-CoV-2 specifically. The reported efficacy of favipiravir, lopinavir/ritonavir and remdesivir was found to be either similar or inferior to the natural component from Guduchi. A previous retrospective study^21^ also found that Guduchi Ghan Vati was effective in viral clearance and reducing hospital stay compared to standard care.

Therefore, based on the available preliminary evidence, the first prospective feasibility study was undertaken. We aimed to investigate the safety of Guduchi Ghan Vati in asymptomatic patients with Covid-19 and to establish an initial impression of its efficacy.

## Methods

In urgent response to the accelerating outbreak of Covid-19 in India, the present study was conducted at Covid Care Centre, Jodhpur, Rajasthan, India, as a nonrandomized, single arm trial. The study was approved by the Institutional Review Board, who independently reviewed the safety of the drug administered. The study complied with the Good Clinical Practice guidelines, Declaration of Helsinki, and Research regulatory requirements of Indian Council of Medical Research along with AYUSH recommended guidelines for patient care with Ayurveda.

Under the principal investigator (AK), the team of investigators (GP, SS, VG) was mainly responsible for designing, initiating and managing trials with the local Jodhpur Covid-19 care center in Rajasthan, India. Aarogyam (UK) as a collaborative technical partner assisted in the study compilation, data management and statistical analyses. Data were recorded by physicians mainly responsible for patient care in hospitals. The research assistant then entered confirmed data, masking the patient identifications and any contact details, into the Medcalc statistical software database for statistical analyses.

Independent of the study team, the Data & Safety Monitoring Board was established, which monitored the progress of the study and reviewed the safety and efficacy data while the trial was ongoing. Committee members represented multidisciplinary experts, including Covid Care physicians, Ayurveda Physicians, and Biostatistician. The committee agreed to share the report of the trial as a priority to support better clinical management of Covid-19 and facilitate future trials.

### Patients

This study enrolled 46 patients who were admitted between May 1, 2020 and May 31, 2020 in the government of Rajasthan designated the Covid-19 treatment centre. There was no placebo used, and the drug was not masked.

All hospitalized cases above 18 years of age, diagnosed with Covid-19 and who are asymptomatic at the time of admission were included in the study. Patients were excluded if they had severe vomiting that would affect oral administration of medicine difficult, respiratory failure or requiring mechanical ventilation, patients having alanine transaminase (ALT) or aspartate transaminase (AST) > 5 times the upper range of normal limits, patients with Covid-19 in critical condition or ARDS or NIAD 8 –point ordinal score-2 (hospitalized, on invasive mechanical ventilation or extracorporeal membrane oxygenation, combined organ failure requiring ICU monitoring, patients with uncontrolled diabetes mellitus, (HbA1c more than 8.0), malignant, chronic renal failure or those on immunosuppressive medication. Eligible patients were given study details and provided an information sheet on Ayurveda medicine along with trial information. Written informed consent was obtained from each participant. Participants were free to withdraw from the study willingly at any stage. All patients were managed with WHO and ICMR clinical practice guidelines for Covid-19. Teams of laboratory technicians who performed clinical measurements were unaware of treatment information.

### Intervention

Guduchi Ghana is a unique Ayuvedic classical preparation prepared from aqueous extracts of Tinospora cordifolia stem. For the purpose of the present study, the VaLJi (tablet) form was prepared in the Pharmacy, University College of Ayurveda, Jodhour, India following the standard procedure. Guduchi Ghan Vati was orally administered 2 tablets (1000 mg) twice daily for 2 weeks.

### Outcome measures

The primary end point was virologic clearance at day 7 post-treatment. Secondary outcomes were clinical follow-up (day 14) (body temperature, clinical symptoms, hospital stay and complications) and the occurrence of any side effects. Outcome measures were collected at baseline, day 3, day 7 and day 14. In addition to SARS-CoV-2 testing, patients were assessed for vital signs and laboratory tests at hospital admission and follow-up at day 14. Records of Guduchi administration, side effects and adverse events were reviewed daily to ensure patient safety.

### Statistical analysis and sample size calculation

This study was taken as a pilot study for asymptomatic patients only. Assuming a 50% efficacy of Guduchi Ghan Vati in reducing the viral load at day 7, an 85% power, a type I error rate of 5%, and 20% dropouts, we calculated that a total of 46 Covid-19 patients would be required for the analysis (Fleiss with CC).

Statistical differences were evaluated by Pearson’s chi-square or Fisher’s exact tests as categorical variables, as appropriate. Means of quantitative data were compared using Student’s t-test. Analyses were performed in MedCalc, version 15.0 (MedCalc Software, Ostend, Belgium).

## Results

Of 92 screened Covid-confirmed patients, 46 asymptomatic patients were included in the study, and 40 completed the 14-day follow-up. The total dropout rate was 13.04%), and all 14 days of drug compliance was 100%. The median participant age was 31 years, and 80% were male. None of the patients had pre-existing conditions. Table 1 describes the main clinical characteristics at baseline. At admission, none of the patients had any symptoms or normal vital and laboratory tests except an elevated erythrocyte sedimentation rate. The interval from the first day of positive nucleic acid tests to the first day of continuous negative tests was 4.82 mean days (in usual setting 14 to 21 days), and 38 (95%) had a negative test on D-7 who were discharged from the hospital on day 9. Two patients had negative results on follow-up D-14. (Table 2)

**Table 1:**
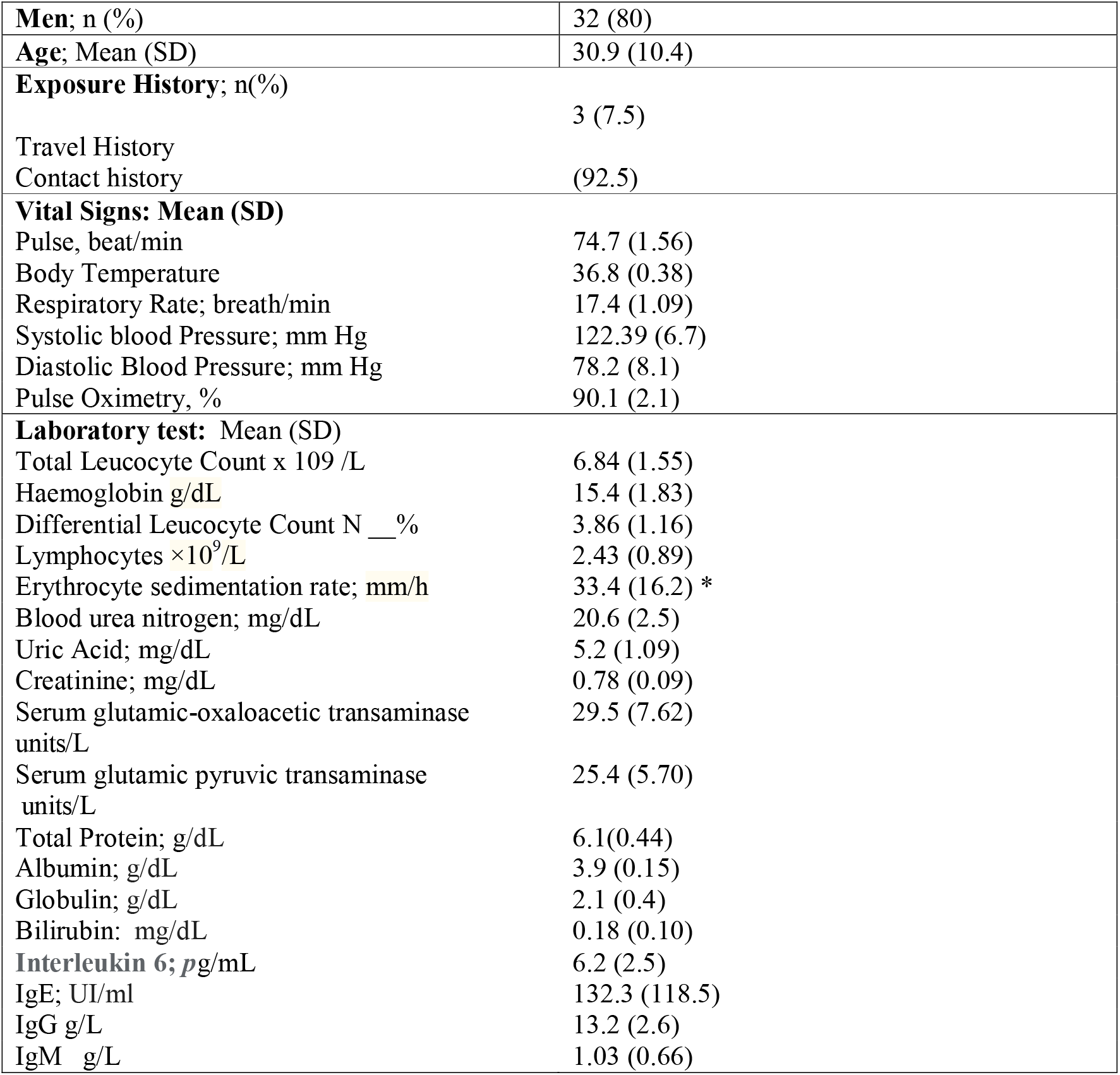
Clinical characteristics at baseline (n=40)

**Table 2:** Proportion with Virological Cure by day in Covid-19 asymptomatic patients treated with Guduchi Ghan Vati.

| Post inclusion | Number of Patients (%) | P value |
| --- | --- | --- |
| Day-3 | 16 (32.5) | <0.0001 |
| Day-7 | 38 (95) | <0.0001 |
| Day-14 | 40 (100) | <0.0001 |

The proportion of patients with negative PCR results in nasopharyngeal samples significant Increased at D-3 (95% CI difference from baseline 17.3039% to 47.9823%; p<0.0001), D-7 (95% CI difference from baseline, 80.5392% to 98.6179%; p<0.0001) and D-14 (95% CI difference from baseline, 87.6084% to 100.0000% p<0.0001).

On the follow-up at D-14, lab tests were repeated, and compared to baseline, ESR levels were reduced significantly (95% CI difference 8.499-19.901; p<0.0001) and were within the normal range along with the rest of the blood tests. There were no adverse events or side effects reported. Ayurveda was safe to use in all study participants.

## Discussion

Taken together, the present study highlights the efficacy and safety of Guduchi Ghan Vati, an Ayurvedic preparation, in the treatment of asymptomatic Covid-19. The results suggest a promising role of Guduchi Ghan Vati in terms of virologic cure with no side effects. Notably, young adults having no pre-existing medical condition might have influenced faster recovery. However, comparing the median duration of viral shedding reported in the previous study was 19 d (IQR, 15–26 d), whereas with Guduchi Ghan Vati, it was 5 d (IQR, 3-11 d).

Major limitations of the study should be considered. First, properly designed controlled trials are urgently required to confirm the benefits of Ayurveda treatment. Second, without any pre-existing symptoms or medical conditions, in the healthy asymptomatic Covid-19 confirmed case, the role of Ayurveda intervention needs to be evaluated with caution.

Finally, background therapies, including support interventions previously taken by participants, were not protocolled. Since the early outbreak, prophylactic support therapies (herbal supplements, Ayurveda, Homeopathy) have been recommended by the Government of India and are widely used by people.

In conclusion, in asymptomatic Covid-19 confirmed cases, Ayurveda intervention can be considered a safe option. The results highlight the importance of curbing infection, which could limit the transmission of the virus to other people to curb the spread of Covid-19.

## Data Availability

Data can be available on request.

## Acknowledgement

We would like to thank the Ministry of AYUSH, Govt of India & Central Council for Research in Ayurvedic Sciences, Ministry of AYUSH, Government of India, New Delhi for their technical support, District & Health administration of Jodhpur for providing necessary permissions, Mr Arun Purohit, Registrar & Mr Tulasi Das Sharma FA for supporting in making necessary arrangements for the study and all residents of DSRR University, Jodhpur for their active involvement in caring for all the study patients.

This study was supported by the Aarogyam (UK) as part of the Research Collaboration. We would like to thank Parashar Sharma (Samta Ayurveda Prakoshtha, India) and Dr Jaydeep Joshi (Aarogyam UK) for their support. Vishwes Kulkarni, University of Warwick, UK, acknowledged data analysis support.

